# Predicting bipolar disorder incidence in young adults using gradient boosting: a 5-year follow-up study

**DOI:** 10.1101/2023.03.31.22282507

**Authors:** Bruno Braga Montezano, Vanessa Gnielka, Augusto Ossamu Shintani, Kyara Rodrigues de Aguiar, Thiago Henrique Roza, Taiane de Azevedo Cardoso, Luciano Dias de Mattos Souza, Fernanda Pedrotti Moreira, Ricardo Azevedo da Silva, Thaíse Campos Mondin, Karen Jansen, Ives Cavalcante Passos

**Author notes:** Corresponding author *Email address:* (Bruno Braga Montezano).

## Abstract

This study aimed to develop a classification model predicting incident bipolar disorder (BD) cases in young adults within a 5-year interval, using sociodemographic and clinical features from a large cohort study. We analyzed 1,091 individuals without BD, aged 18 to 24 years at baseline, and used the XGBoost algorithm with feature selection and oversampling methods. Forty-nine individuals (4.49%) received a BD diagnosis five years later. The best model had an acceptable performance (test AUC: 0.786, 95% CI: 0.686, 0.887) and included ten features: feeling of worthlessness, sadness, current depressive episode, selfreported stress, self-confidence, lifetime cocaine use, socioeconomic status, sex frequency, romantic relationship, and tachylalia. We performed a permutation test with 10,000 permutations that showed the AUC from the built model is significantly better than random classifiers. The results provide insights into BD as a latent phenomenon, as depression is its typical initial manifestation. Future studies could monitor subjects during other developmental stages and investigate risk populations to improve BD characterization. Furthermore, the usage of digital health data, biological, and neuropsychological information and also neuroimaging can help in the rise of new predictive models.

## 1. Introduction

Bipolar disorder is a chronic psychiatric condition associated with high morbidity and mortality (McIntyre et al., 2020). The global lifetime prevalence of bipolar disorder is approximately 2.4%, being 0.6% for bipolar I disorder, 0.4% for bipolar II disorder and 1.4% for individuals with subthreshold presentations (Merikangas et al., 2011). Previous investigations describe that these patients present a significant reduction of life expectancy of about 10 years relative to the general population (Kessing et al., 2015). Cardiovascular disease is the main factor associated with premature mortality in bipolar disorder; nonetheless, deaths by suicide are more commonly reported in bipolar disorder than in other mental health conditions, with these patients presenting a twenty to thirty times higher chance of dying by suicide (McIntyre et al., 2020; Plans et al., 2019; Kessing et al., 2015). In addition, bipolar disorder patients present significant functional and psychosocial impairment, also representing an important economic cost (McIntyre et al., 2020). For instance, evidence from the United States described that the total costs associated with bipolar I disorder exceeded $200 billion in the year of 2015 (Cloutier et al., 2018).

Even though the majority of the patients with BD present clinical symptoms before the age of 25, there is a significant delay of 6-10 years between the onset of the symptoms and the correct diagnosis (Yatham et al., 2018; Scott & Leboyer, 2011). Furthermore, delayed diagnosis is associated with longer duration of untreated illness, which is ultimately linked to a poorer prognosis in terms of hospitalizations, functioning and recurrence of episodes (Altamura et al., 2015). High rates of psychiatric comorbidity, difficulty in the differential diagnosis, usual onset with depressive symptoms, and reduced help-seeking behavior are some of the reasons for the delay in the proper recognition of bipolar disorder (McIntyre & Calabrese, 2019; McIntyre et al., 2020; Yatham et al., 2018). Nevertheless, the diagnosis of bipolar disorder is eminently clinical, with limited evidence to support the use of neuroimaging or laboratory biomarkers during clinical investigation (McIntyre et al., 2020).

Taking into account this context, the rise of the concept of precision psychiatry, with the use of big data and machine learning tools represents a promise, which may ultimately bring a revolution in terms of diagnosis, treatment selection and prognosis in the field of mental health (Fernandes et al., 2017; Passos et al., 2016). To this date several studies have explored the use of these techniques in bipolar disorder, based on distinct data sources (including neuroimaging, clinical and sociodemographic data, peripheral biomarkers, neuropsychological tests, genetics, among others), with the majority of these models being focused on classification tasks that help in the differential diagnosis between bipolar disorder and other psychiatric conditions such as schizophrenia, major depression and healthy individuals (Librenza-Garcia et al., 2017; Passos et al., 2019). Nevertheless, most of these studies present modest classification performances, are based on small and clinical samples, originary from cross-sectional procedures of data collection, or present short periods of follow-up (Librenza-Garcia et al., 2017). All these limitations may compromise the generalizability and the translation of the results of such investigations to clinical and public health settings (Passos et al., 2019).

Thus, considering these gaps, the present study aims to create a binary classification model capable of predicting incident cases of bipolar disorder in a 5-year interval through sociodemographic and clinical features in a sample of young adults, from a large and population-based cohort study.

## 2. Methods

### 2.1. Participants

This was a prospective cohort study that collected sociodemographic and clinical information from a population-based sample of young adults aged between 18 and 24 years, living in the urban area of the city of Pelotas, located in southern Brazil. The first phase took place between 2007 and 2009, and the sample was selected through cluster sampling, considering eighty-nine randomly selected census-based sectors from 448 total sectors (Brazilian Institute of Geography and Statistics, 2010).

The following inclusion criteria were considered at baseline: (1) age between 18 and 24 years old; (2) live in the urban area. Severe cognitive disability (assessed through clinical judgement) that could cause difficulties in understanding study instruments was considered the only exclusion criteria. All eligible subjects (*n* = 1762) were invited to participate, of which 1560 accepted and consented to participate. Trained interviewers conducted a face-to-face interview at the participants’ homes, so that data confidentiality was ensured. Data were collected through printed paper questionnaires with research instruments and diagnostic criteria for mental disorders.

The follow-up occurred from 2012 to 2014, that is, an average interval of five years after the first assessment. The participants from baseline (*n* = 1560) were invited for a second data collection. All interviewers met weekly to discuss the assessments, focusing on those who were uncertain about the BD diagnosis. In these situations, a psychiatrist was recruited to carry out the reassessment. 1244 individuals were located and consented to be reevaluated (79.7% of retention), and 14 (0.9%) were lost due to death. Since the present study aims to predict BD incidence, subjects who met diagnostic criteria for a lifetime manic or hypomanic episode were excluded. Unlike the baseline, data were collected through tablets using Open Data Kit (ODK), an open-source mobile data collection platform (Hartung et al., 2010). The forms were filled out offline, and the data were later backed up to computers through secure data transfer protocols. This study was approved by the Research Ethics Committee of Universidade CatÓlica de Pelotas under protocol number 2008/118. The subjects who presented any psychiatric diagnosis in the clinical interview were referred for specialized treatment in the local health system. All participants signed a printed informed consent form and could withdraw from the study at any time.

### 2.2. Outcome

The BD diagnosis was built with modules A and D from Mini International Neuropsychiatric Interview 5.0 (MINI), in order to assess current or past depressive episodes and current or past manic or hypomanic episodes, respectively. The BD diagnoses were reassessed in those cases where the diagnosis was questionable. MINI is a short-term diagnostic interview designed for clinical assessment of mental disorders according to Diagnostic and Statistical Manual of Mental Disorders — Fourth Edition (DSM-IV) and ICD-10 criteria (American Psychiatric Association, 1994; World Health Organization, 1993). Despite evaluating several disorders, MINI psychometric properties for the diagnosis of lifetime manic episode (sensitivity: 81.0%; specificity: 94.0%; positive predictive value: 76.0%; negative predictive value: 95.0%) and major depressive episode (sensitivity: 96.0%; specificity: 88.0%; positive predictive value: 87.0%; negative predictive value: 97.0%) are reliable when compared to DSM Structured Clinical Interview (Amorim, 2000).

### 2.3. Predictors

One hundred and ninety features were included in the original dataset before preprocessing steps. These variables include demographic, social, clinical, and environmental characteristics. The following features were included in the modeling pipeline:

a) Sociodemographic and environmental variables: Sex, skin color, age, socioeconomic status (3 levels and 5 levels), current occupation, currently studying, worked for money, has a partner, has a religion, access to psychotherapy, knows someone who attempted suicide or committed suicide, involvement in physical fights, family gun ownership, social support, has divorced parents, has any deceased parents, has someone close by already deceased, individual and family stress problems, lives with parents, family suicide attempts, seat belt wearing, helmet use when riding a motorcycle, suffered an accident that led to an emergency room, drove or took a ride with a drunk driver.
b) Substance use variables: Indicative of substance abuse or dependence (tobacco, alcohol, cannabis, cocaine, crack, amphetamines, inhalants, sedatives, hallucinogens, opioids, illicits, any other substances) assessed by Alcohol, Smoking and Substance Involvement Screening Test (ASSIST) and lifetime use features (same substances cited above), age that first used drugs, injected drugs use, use of medication for stress problems in the last 30 days.
c) Clinical variables: mental disorder diagnoses (anxiety, mood and personality disorders), eating disorders, current suicide risk, serious organic disease, lifetime psychiatrist or psychologist visit, lifetime psychotherapeutic treatment, interrupted treatment.
d) Sex-related variables: age of first sexual intercourse, sexual intercourse in the last week (sex frequency), condom use, alcohol use before sexual intercourse, number of sexual partners, number of pregnancies, lifetime sexual abuse, lifetime sexual intercourse (binary).
e) Psychometric instrument items: Beck Depression Inventory (BDI) [21 items], Hypomania Checklist (HCL-32) [32 items], Social Readjustment Rating Scale (SRRS) [26 items], Beck Scale for Suicide Ideation (BSS) [21 items].

### 2.4. Machine learning analysis

Aiming to predict new cases of bipolar disorder in young adults, using features previously described, we created an ML pipeline to generate a predictive model using supervised learning. We used a vastly used machine learning algorithm for tabular data called tree gradient boosting, implemented through the *XGBoost* library (Chen & Guestrin, 2016) in the R programming language on version 4.2.1 (R Core Team, 2022).

Tree gradient boosting is part of what is called ensemble algorithms — joining many models to make predictions together — in statistical learning methods. Boosting improves this concept by building a sequence of originally weak models into progressively more powerful models. Additionally, in gradient boosting techniques, the gradient of a loss function is used to choose the best approach to improve a weak learner (James et al., 2021). In the context of gradient-boosted trees, weak learners are decision trees.

The following tree boosting hyperparameters were tuned (Kuhn & Vaughan, 2022a; Chen & Guestrin, 2016):

*a) mtry*: Number of predictors that is randomly sampled at each split.

*b) trees*: Number of trees contained in the ensemble.

*c) min n*: Minimum number of observations in a node required for the node to be split further.

*d) tree depth*: Maximum depth (number of splits) of each tree.

*e) loss reduction*: Reduction in the loss function required to split further.

*f) learn rate*: Step size at each iteration while moving toward a loss function optimization.

*g) sample size*: Proportion of the data set used for modeling within an iteration.

When it comes to tabular data, gradient boosting decision trees (GBDT) are seen as the state-of-the-art, reinforced by several competitions in the ML scenario. In addition, a study found that GBDT perform better than deep learning models across multiple tabular datasets, and also requires less hyperparameter tuning (Shwartz-Ziv & Armon, 2021).

The implementation of the data modeling routines was carried out using the *tidymodels* framework (Kuhn & Wickham, 2020). The *tidymodels* is a R metapackage made up of multiple packages that assist in different stages of a machine learning pipeline. In order to split the data and create cross-validation resamples, *rsample* package was used (Silge et al., 2022), *parsnip* was used to access *XGBoost* functions in an unified manner (Kuhn & Vaughan, 2022a), *recipes* for preprocessing functions (Kuhn & Wickham, 2022), *workflows* to bundle the preprocessing, modeling and post-processing routines (Vaughan, 2022), *yardstick* to easily calculate performance measures (Kuhn & Vaughan, 2022b).

In the cross-validation, fifteen hyperparameter combinations were used as candidate parameter sets. The values for each hyperparameter were randomly chosen based on an algorithm that attempts to maximize the determinant of the spatial correlation matrix between coordinates (Santner et al., 2003).

### 2.5. Preprocessing

Before any preprocessing routine was performed, the data was divided into two subsets. A training set, consisting of 70% of the sample, and a test set with the remaining samples (30% of the total samples). The entire test set was isolated until the end of all tuning and validation procedures to build the model, to then be used to simulate the model performance on new data.

Some preprocessing techniques were adopted to clean and tidy data prior to modeling. The following preprocessing steps were applied:

1) Remove all features with more than 10% of missing values.

2) Impute categorical features with mode.

3) Impute numeric features with median.

4) Remove near-zero variance features (few unique values relative to the number of observations and also a ratio of the frequency of the second most common value is large [ratio of 10]).

5) Create dummy variables with *C -* 1 categories from categorical features.

### 2.6. Feature selection

The feature selection process aims to automatically filter variables from the data matrix considering their relevance to the predictive modeling problem. Therefore, we can build more accurate and parsimonious models while, at the same time, saving computational resources through the use of less data in the next model fitting steps.

The Boruta system was implemented for feature selection in the present pipeline. It consists of a random forest based algorithm that iteratively removes features that are statistically less important than random synthetic features (artificial noise). For each iteration, removed variables are prevented from being considered for the next iteration (Kursa et al., 2010). Boruta is considered a wrapper method as it takes into account a subset of variables with different combinations in each iteration.

### 2.7. Class imbalance

Class imbalance is a common problem in classification modeling. It happens when we face a set of examples that presents a given level way more frequently than other. Since most ML classifiers assume data equally distributed, they tend to be more biased towards the majority class, causing bad performance on minority class classification.

This concept is especially important in the context of predicting mental disorders, as subjects who will present the disease will be exposed to greater health risk. Therefore, it is necessary that the classifiers of such outcomes can adequately predict this portion of the population. BD still has the aggravating factor of having a complex prognosis regarding the neuroprogression, which can be worsened by the length of disease (Librenza-Garcia et al., 2021).

For this paper, an algorithm named ROSE (Random Over-Sampling Examples) was used. ROSE is a smoothed-bootstrap-based technique that creates new artificial observations in data in order to minimize or eliminate class imbalance (Menardi & Torelli, 2014). The *themis* and *ROSE* R packages were adopted to implement the previously described algorithm (Hvitfeldt, 2022; Lunardon et al., 2014).

### 2.8. Cross-validation

The cross-validation (CV) process was used to tune *XGBoost* hyperparameters described earlier. We used the *k*-fold cross-validation technique with 5 folds repeated five times. In order to optimize the hyperparameter combinations, we used a racing method proposed by Kuhn (2014). It consists in calculating the area under the receiver operating characteristic (ROC) curve for each parameter set across validation folds. After evaluating the parameter combinations for three resamples, a repeated measure ANOVA model is fitted. The combinations that are statistically different (based on *α* level for one-sided confidence interval of 5%) from the best setting are excluded from further validation procedures. The ANOVA racing method was implemented via *finetune* R package (Kuhn, 2022).

In Figure 1, the cross-validation procedure can be visualized inside the orange area. For each fold, the Boruta and ROSE algorithms were applied just in training folds, leaving the testing fold untouched in order to properly estimate model error. A maximum of 25 runs (5-fold CV repeated up to five times) was considered for hyperparameter tuning. At the end, the remaining models of the ANOVA racing process were evaluated, and the model with the highest validation AUC was chosen to be tested in testing set from the initial data split.

**Figure 1:**
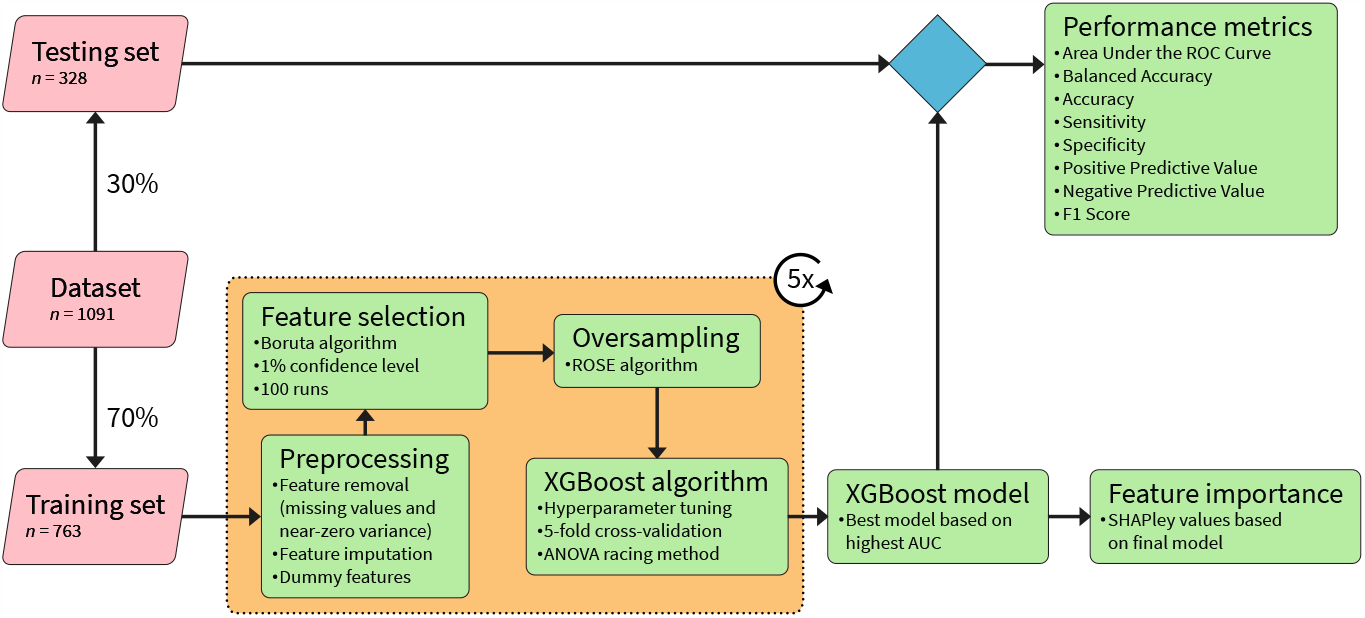
Machine learning pipeline flowchart. The figure shows data splitting, preprocessing routine, feature selection, cross-validation, model fitting, model assessment and feature importance steps using *XGBoost* algorithm.

### 2.9. Performance measures

Aiming to evaluate the performance of the algorithm, some evaluation metrics were used. Firstly, the area under the receiver operating characteristic (ROC) curve (AUC) was used to diagnose the classifier ability to predict correctly across multiple discrimination thresholds (Fawcett, 2006).

Sensitivity and specificity were also used to assess model ability to correctly detect subjects with BD who have the disorder, and correctly detect subjects that did not present BD who actually does not have BD, respectively (Yerushalmy, 1947). Positive and negative predictive values (PPV and NPV) show the proportion of positive and negative predictions that are truly positive or negative, correspondingly.

Accuracy measures the proportion of correct predictions among the total number of observations evaluated (Metz, 1978). In order to take into account the class imbalance previously described, we also used balanced accuracy as it inputs both sensitivity and specificity into its formula:

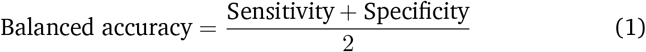

The F1 score is defined as the harmonic mean of PPV and sensitivity (Equation 2). This metric is able to demonstrate another model accuracy measure, being more robust in class imbalance scenarios.

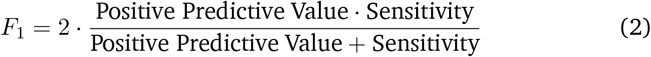

### 2.10. Model interpretability

In order to make model predictions more interpretable, we used SHAPley values. SHAPley values shows how to fairly distribute the total output among all features. Beyond that, SHAP (SHapley Additive exPlanations) allow for explanation on individual predictions (Lundberg & Lee, 2017). In the present study, the SHAPley values were obtained using the R package *SHAPforxgboost* (Liu & Just, 2021). This package provides functions to create SHAP-related visualizations from a *XGBoost* model object.

In addition to the use of feature importance visualization, partial dependence plots (PDP) were also employed. They are able to show the marginal effect a feature has on the predicted outcome of a machine learning model (Friedman, 2001). The PDP were built with the *pdp* (Greenwell, 2017) and *SHAPforxgboost* (Liu & Just, 2021) R packages, along with the *ggplot2* (Wickham, 2016) and *patchwork* (Pedersen, 2020) packages for plot composition.

## 3. Results

The present study aimed to create a model to predict bipolar disorder onset on young adults based on 5-year follow-up data. We assessed 1,091 subjects at follow-up interview who had no current or past episode of mania or hypomania at the first assessment. Of these, 4.49% (*n* = 49) young adults received a diagnosis of BD five years later. Descriptive tables of demographic features at baseline are presented in Table 1. Absolute and relative frequency of missing values in each feature are described in Table 2. Table 3 shows the selected hyperparameter set from the cross-validation.

**Table 1:** Sociodemographic features measured at baseline grouped by bipolar disorder diagnosis in the follow-up.

| Features | With BD ($n = 49$) | Without BD ($n = 1042$) | Overall ($n = 1091$) | $p$-value |
| --- | --- | --- | --- | --- |
| **Sex** |  |  |  | 0.302 |
| Male | 16 (32.7%) | 457 (43.9%) | 473 (43.4%) |  |
| Female | 33 (67.3%) | 585 (56.1%) | 618 (56.6%) |  |
| **Age** |  |  |  | 0.353 |
| Mean (SD) | 20.9 (2.28) | 20.5 (2.10) | 20.5 (2.11) |  |
| Median [Min, Max] | 21.0 [17.0, 25.0] | 20.0 [16.0, 26.0] | 20.0 [16.0, 26.0] |  |
| Missing | 0 (0%) | 8 (0.8%) | 8 (0.7%) |  |
| **Socioeconomic status** |  |  |  | 0.635 |
| Upper | 14 (28.6%) | 401 (38.5%) | 415 (38.0%) |  |
| Middle | 26 (53.1%) | 509 (48.8%) | 535 (49.0%) |  |
| Lower | 9 (18.4%) | 132 (12.7%) | 141 (12.9%) |  |
| **Skin color** |  |  |  | 0.800 |
| White | 34 (69.4%) | 765 (73.4%) | 799 (73.2%) |  |
| Non-white | 15 (30.6%) | 273 (26.2%) | 288 (26.4%) |  |
| Missing | 0 (0%) | 4 (0.4%) | 4 (0.4%) |  |
| **Currently working** |  |  |  | 0.965 |
| No | 8 (16.3%) | 245 (23.5%) | 253 (23.2%) |  |
| Yes | 14 (28.6%) | 380 (36.5%) | 394 (36.1%) |  |
| Missing | 27 (55.1%) | 417 (40.0%) | 444 (40.7%) |  |
| **Has a partner** |  |  |  | 0.953 |
| No | 13 (26.5%) | 247 (23.7%) | 260 (23.8%) |  |
| Yes | 33 (67.3%) | 696 (66.8%) | 729 (66.8%) |  |
| Missing | 3 (6.1%) | 99 (9.5%) | 102 (9.3%) |  |

The $p$ -values were calculated from $t$ -tests for numeric features and $\chi^2$ -squared tests for categorical features.

**Table 2:** Missing values of each feature in the entire dataset (*n* = 1091), before data splitting procedure. Only features with at least one missing value are present in the table. All features were collected at baseline.

| Feature | Number of missing values | % of missing values |
| --- | --- | --- |
| Age | 8 | 0.73 |
| Skin color | 4 | 0.37 |
| Currently working (last year) | 444 | 40.70 |
| Deceased parents | 1 | 0.09 |
| Has a religion | 1 | 0.09 |
| Psychiatric hospitalization | 2 | 0.18 |
| Stress problems | 2 | 0.18 |
| Psychiatric medication use | 8 | 0.73 |
| Maternal stress problems | 1 | 0.09 |
| Paternal stress problems | 1 | 0.09 |
| Sibling stress problems | 2 | 0.18 |
| Grandparents stress problems | 1 | 0.09 |
| Partner suicide attempt | 753 | 69.02 |
| Children suicide attempt | 785 | 71.95 |
| Paternal suicide attempt | 725 | 66.45 |
| Maternal suicide attempt | 718 | 65.81 |
| Sibling suicide attempt | 725 | 66.45 |
| Friend suicide attempt | 712 | 65.26 |
| Lifetime sexual intercourse | 8 | 0.73 |
| Worked for money | 6 | 0.55 |

Table 2 continued from previous page
| Feature | Number of missing values | % of missing values |
| --- | --- | --- |
| Has a significant disease | 2 | 0.18 |
| Has ever visited a psychiatrist of psychologist | 3 | 0.27 |
| Has ever been treated by a psychiatrist or psychologist | 107 | 9.81 |
| BSS item 1 | 4 | 0.37 |
| BSS item 2 | 2 | 0.18 |
| BSS item 3 | 4 | 0.37 |
| BSS item 4 | 1 | 0.09 |
| BSS item 5 | 1 | 0.09 |
| BSS item 6 | 1 | 0.09 |
| BSS item 7 | 1 | 0.09 |
| BSS item 8 | 1 | 0.09 |
| BSS item 9 | 1 | 0.09 |
| BSS item 10 | 1 | 0.09 |
| BSS item 11 | 1 | 0.09 |
| BSS item 12 | 1 | 0.09 |
| BSS item 13 | 1 | 0.09 |
| BSS item 14 | 1 | 0.09 |
| BSS item 15 | 1 | 0.09 |
| BSS item 16 | 1 | 0.09 |
| BSS item 17 | 1 | 0.09 |
| BSS item 18 | 1 | 0.09 |

Table 2 continued from previous page
| Feature | Number of missing values | % of missing values |
| --- | --- | --- |
| BSS item 19 | 1 | 0.09 |
| BSS item 20 | 3 | 0.27 |
| BSS item 21 | 1057 | 96.88 |
| Knows someone who tried suicide | 1 | 0.09 |
| BDI item 1 (sadness) | 1 | 0.09 |
| BDI item 2 (hopelessness about the future) | 1 | 0.09 |
| BDI item 3 (feeling like a failure) | 1 | 0.09 |
| BDI item 4 (anhedonia) | 1 | 0.09 |
| BDI item 5 (sense of guilt) | 1 | 0.09 |
| BDI item 6 (feel that you are being punished) | 1 | 0.09 |
| BDI item 7 (self-hatred) | 1 | 0.09 |
| BDI item 8 (blame yourself for everything) | 1 | 0.09 |
| BDI item 9 (death thoughts) | 1 | 0.09 |
| BDI item 10 (excessive crying) | 1 | 0.09 |
| BDI item 11 (irritability) | 1 | 0.09 |
| BDI item 12 (loss of interest in personal relationships) | 1 | 0.09 |
| BDI item 13 (impaired decision making) | 1 | 0.09 |
| BDI item 14 (self-image) | 1 | 0.09 |
| BDI item 15 (occupational performance) | 1 | 0.09 |
| BDI item 16 (sleep) | 1 | 0.09 |
| BDI item 17 (fatigue) | 1 | 0.09 |

Table 2 continued from previous page
| Feature | Number of missing values | % of missing values |
| --- | --- | --- |
| BDI item 18 (loss of appetite) | 1 | 0.09 |
| BDI item 19 (weight loss) | 1 | 0.09 |
| BDI item 20 (physical health concern) | 1 | 0.09 |
| BDI item 21 (loss of sexual interest) | 1 | 0.09 |
| Age that first used drugs | 139 | 12.74 |
| Age of first sexual intercourse | 111 | 10.17 |
| Sex frequency (sexual intercourse in the last week) | 106 | 9.72 |
| Condom use during last sexual intercourse | 101 | 9.26 |
| Alcohol use before the last sexual intercourse | 101 | 9.26 |
| Has a partner | 102 | 9.35 |
| Number of people you had sex with in the last year | 138 | 12.65 |
| Number of times you have gotten or made someone pregnant | 114 | 10.45 |
| Have you experienced forced sex? | 405 | 37.12 |
| Current anorexia nervosa | 1 | 0.09 |
| Antisocial personality disorder | 2 | 0.18 |
| Do you have access to psychotherapy (public or private healthcare)? | 964 | 88.36 |
| Current melancholic depressive episode | 1 | 0.09 |
| Past depressive episode | 1 | 0.09 |

**Table 3:** Hyperparameter set chosen for final model based on highest area under the receiver operating characteristic curve in cross-validation routine.

| Hyperparameter | Value on final model |
| --- | --- |
| *mtry* | 9 |
| *trees* | 1429 |
| *min_n* | 10 |
| *tree_depth* | 14 |
| *learn_rate* | 0.0017 |
| *loss_reduction* | 0.6120 |
| *sample_size* | 0.9075 |

*XGBoost* showed an acceptable performance predicting BD five years before the diagnosis with a test set AUC of 0.786 [95% CI: 0.686, 0.887] (Figure 2). The other performance metrics using a cut-off of 0.5 for class decision boundary^1^ can be seen in Table 4.

**Table 4:** Performance metrics from the *XGBoost* model applied on testing set using 0.5 as threshold for positive classification. Area under the receiver operating characteristic curve: 0.786.

| Performance metrics | *XGBoost* on test set |
| --- | --- |
| Sensitivity (recall) | 0.375 |
| Specificity | 0.920 |
| PPV | 0.194 |
| NPV | 0.966 |
| Balanced accuracy | 0.647 |
| Accuracy | 0.893 |
| F1-score | 0.255 |
| Positive predictive value or precision (PPV); Negative predictive value (NPV). |  |

**Figure 2:**
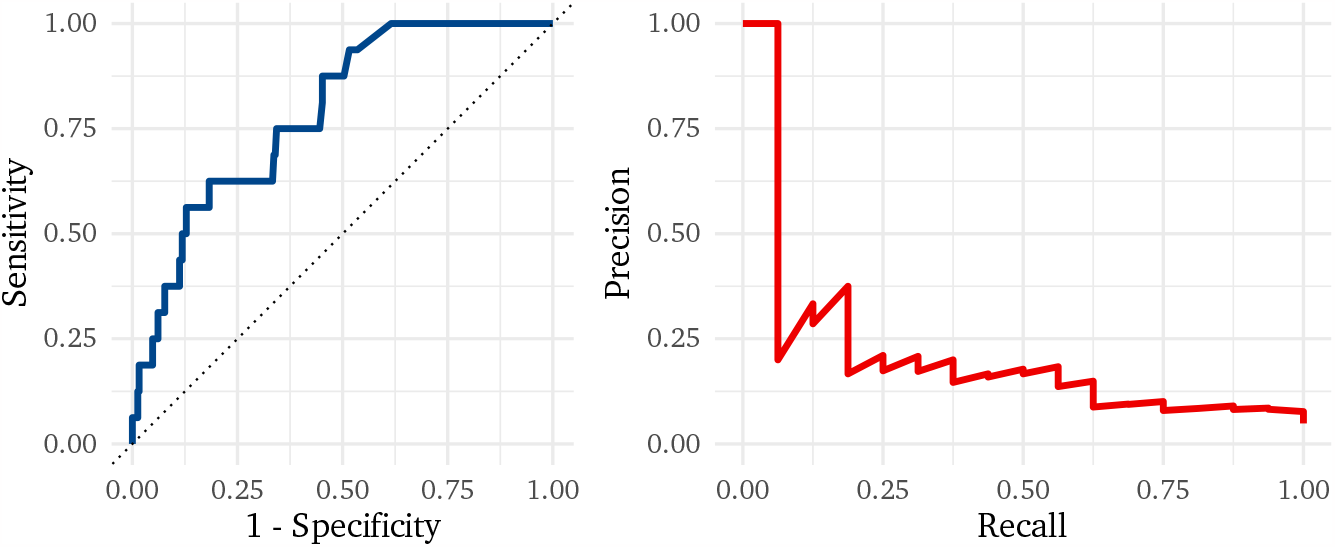
Receiver operating characteristic (ROC) curve and precision recall (PR) curve of the final model fitted on the training set with best parameter combination from cross-validation step, assessed on test set, with an area under the ROC curve of 0.786 and area under the PR curve of 0.208.

The six most relevant baseline features in BD prediction were feeling like a failure (BDI item 3), sadness (BDI item 1), current depressive episode, selfreported stress problems, self-confidence (HCL-32 item 3) and lifetime cocaine use. Feature importance can be seen in more detail in Figure 3. Given the importance of interpreting the model trajectory to a given prediction, to visualize the influence of each feature on the prediction of a specific sample, a force plot was built. The SHAPley values for each training sample is shown in Figure 4. Partial dependence plots can be seen in Figure 5.

**Figure 3:**
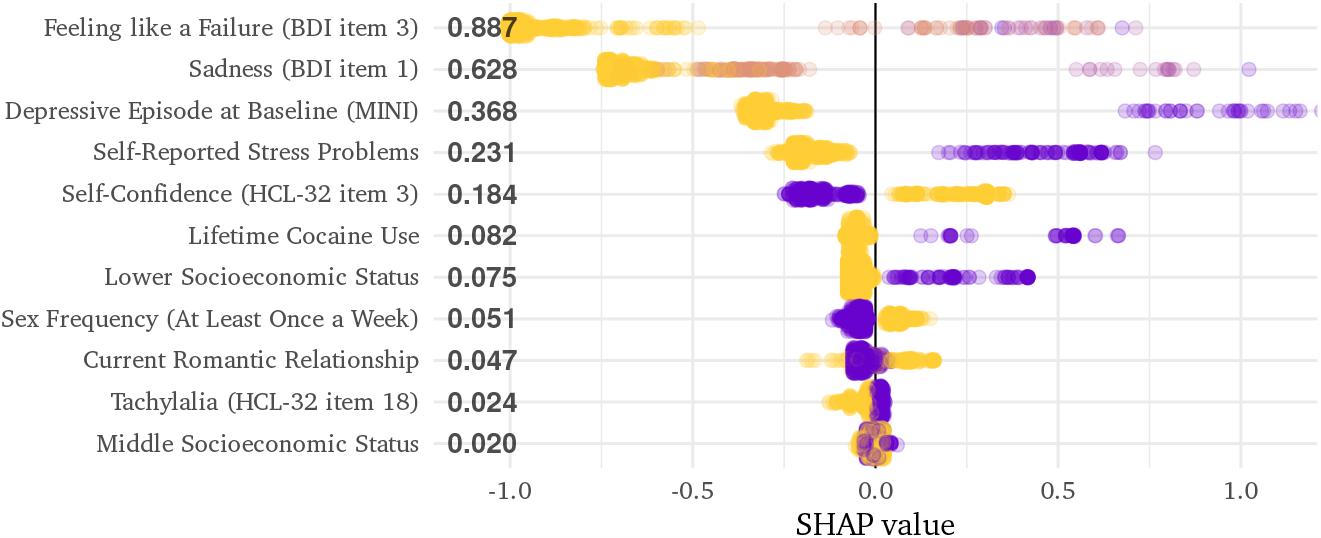
SHAPley values for each feature included in the final model. Each y-axis tick represents a feature, sorted by the highest absolute contribution across all observations, regardless of the direction of the association. Each dot represents a participant in the training set (*n* = 763). Observations with SHAPley values lower than zero behaved as protective factors, otherwise they were risk factors. The fill color represents the value of the variable for a given individual (purple corresponds to higher values, and yellow corresponds to lower values).

**Figure 4:**
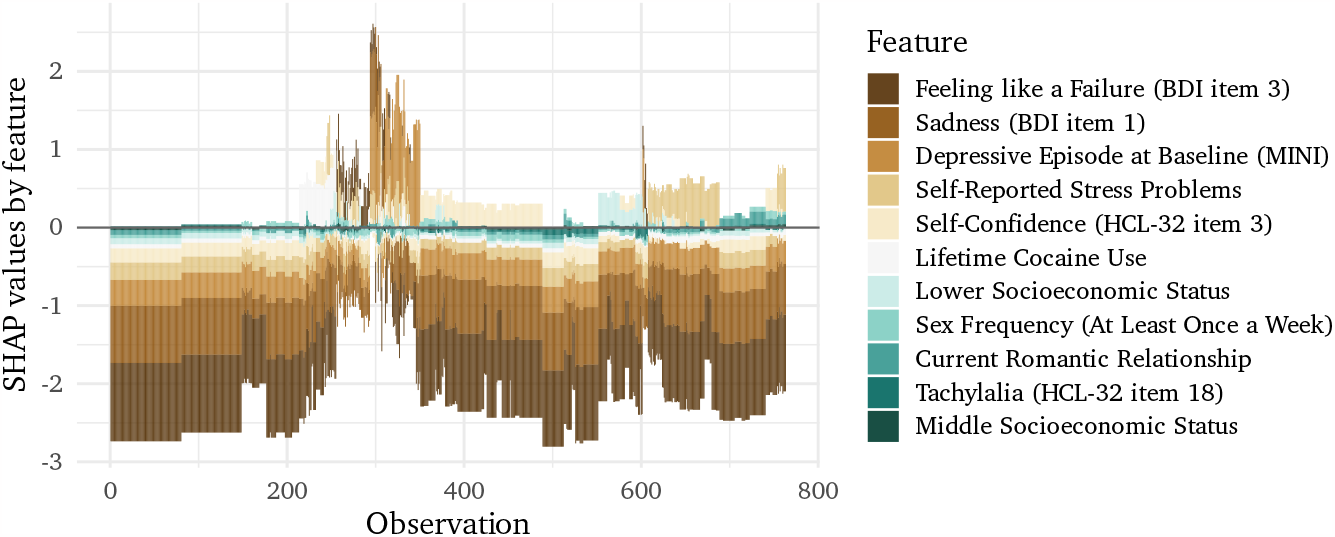
SHAPley force plot. The y-axis demonstrates the influence of each feature on current prediction based on SHAPLey values. The x-axis represents all samples used to train the final model. The whole training set (*n* = 763) is presented.

**Figure 5:**
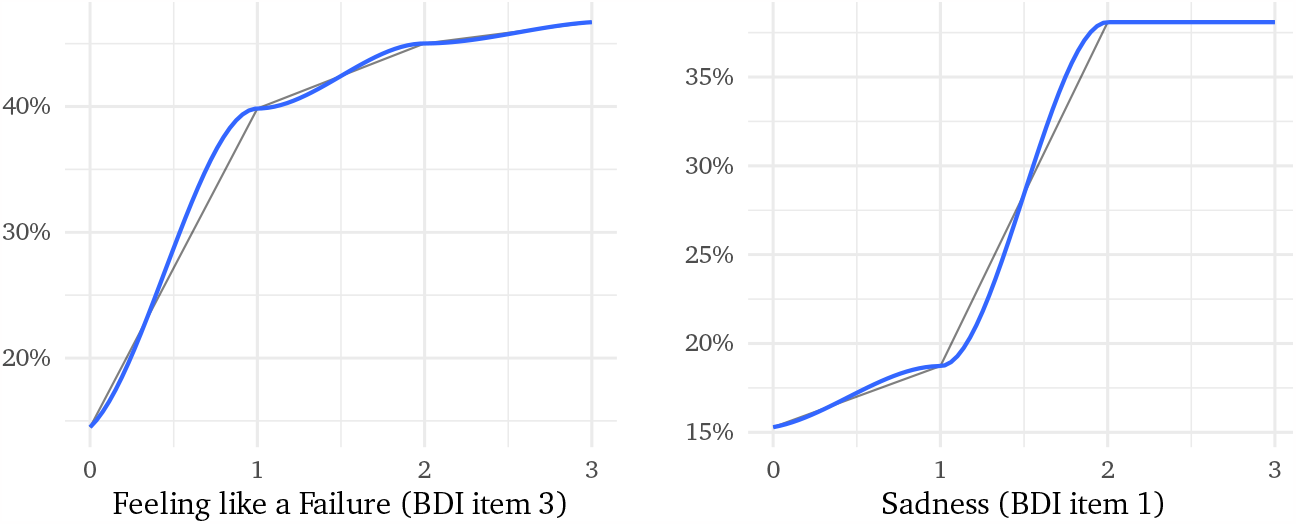
Partial dependence plots for continuous depression-related features. It shows the average trend of each feature. Other variables are held constant. The plots show an upward trend, which indicates that the higher the values of the variables of feeling like a failure and sadness, the higher the predicted probabilities for developing bipolar disorder after five years. The blue line indicates a regression line using the LOESS (locally weighted polynomial regression) method.

In addition to the main pipeline, 1,000 different random training and testing splits were sampled in order to fit the final model. The estimates can be visualized in Figure 6. In this way, an adequate AUC can be seen in the model performance — within the estimated confidence intervals — including a robustness in the predictive power shown through the resamples. Along with the random splits, we also performed a permutation test as proposed by Fisher (1935) to compare the distribution of ROC AUC performance of random rearrangements of the outcome with the original test data using 10,000 permutations. We observed statistical difference between the original and permuted models (*p<*0.001). The distribution of the permuted AUCs is available in Figure 7. This result shows that our model predictions for BD incidence after the five years are more accurate than random classifiers.

**Figure 6:**
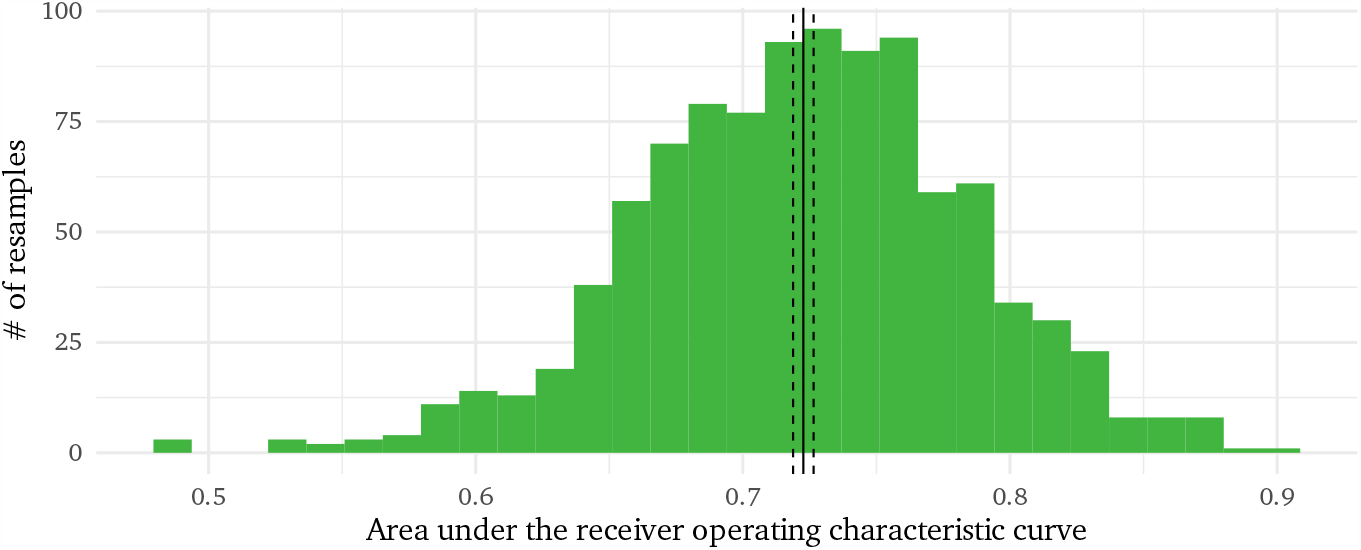
Histogram of the area under the receiver operating characteristic curve (AUC) based on 1,000 random training and testing data splits. The AUC mean and 95% CI found were 0.723 [0.719, 0.726]. This analysis is able to demonstrate the predictive performance robustness of the selected boosting model.

**Figure 7:**
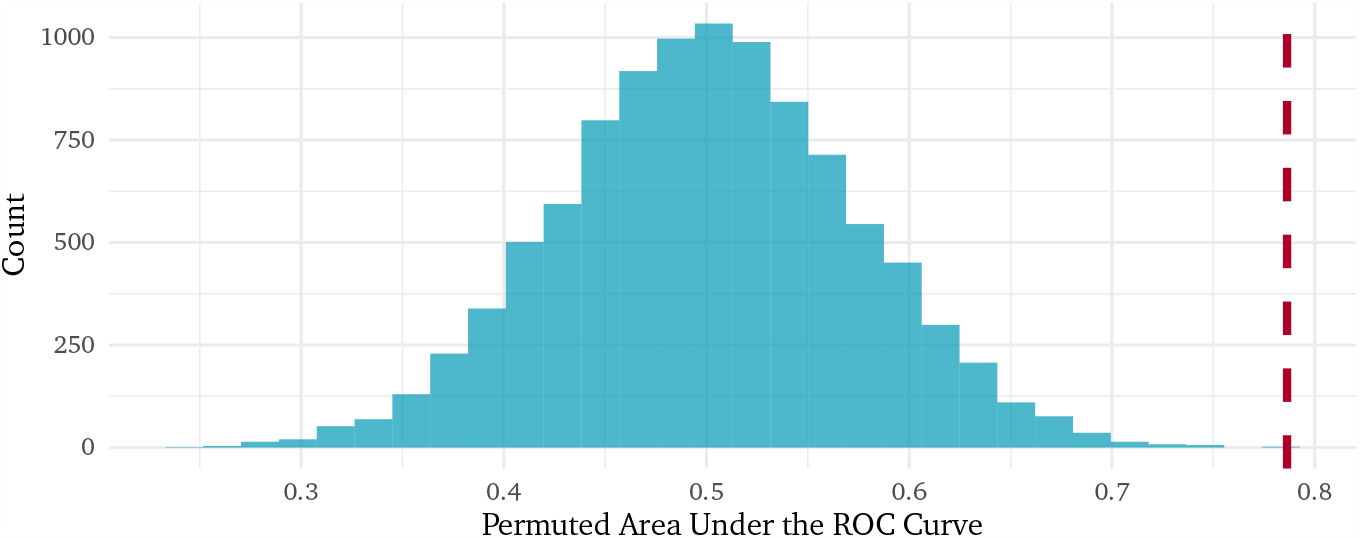
Distribution of the areas under the receiver operating characteristic curves (AUC) from the permutation test with 10,000 rearrangements. The red line indicates the AUC on the original test set (AUC = 0.786).

## 4. Discussion

We proposed to create a model capable of predicting BD in a 5-year interval with acceptable classification performance. Our final model performed with good metrics (AUC: 78.6%), suggesting good predictive capacity. To the best of our knowledge, this is the second Brazilian study to investigate the prediction of bipolar disorder incidence. A previous study investigated the development of a prediction model, with the use of elastic net algorithms, to identify participants who would develop bipolar disorder over the follow-up, in a large community birth cohort, from the city of Pelotas in Brazil (Rabelo-da Ponte et al., 2020). According to the results of this investigation, the model with the best performance (AUC of 0.82) predicted bipolar disorder at the age of 22 years, using clinical and sociodemographic data from the age of 18 years (Rabelo-da Ponte et al., 2020). A recent systematic review on clinical prediction models in psychiatry pointed to several aspects that predictive models could improve, such as overfitting prevention, generalizability and clinical utility (Meehan et al., 2022). The present paper used a larger sample than most studies to predict BD with statistical learning, despite having a low value of events per variable (EPV) of approximately 5.8.

The current study corroborates previous findings in which depressive symptoms would be one of the main predictors for BD conversion (Hafeman et al., 2017; Perich et al., 2015). Notably, the three primary factors found by the prediction model for BD developed in this paper are correlated constructs linked to depression: failure feeling, sadness and current depressive episode. This finding suggests that these factors could be prodromal symptoms of the disorder (Faedda et al., 2019; Van Meter et al., 2016), or even evidence of genetic predisposition to emotional distress (Smeland et al., 2018). It also reinforces the perspective of BD as a worsening trajectory, and the first mood episode as a milestone signaling for a complex disorder onset (Duffy et al., 2014). In the vast majority of cases, the first mood episode of a patient with BD is a depressive one, often years prior to a manic episode (Mesman et al., 2017; Duffy et al., 2014; Mesman et al., 2013), which turns the model also useful to ease the differential diagnosis between unipolar and bipolar depression, because together with other predictors, it is possible to verify whether there is a greater chance that a given current depressive episode is from a unipolar or a bipolar clinical condition.

Lifetime cocaine use is another major predictor evaluated in our study. Several studies have investigated the role of substance use in general and its association with the development of mood disorders. A cross-sectional study conducted in 2013 identified subsequent mood disorders developed in individuals with primary substance use disorder (SUD), and the average time between SUD onset and mood disorder was 11 years (Kenneson et al., 2013). The odds of developing bipolar disorder were particularly high among individuals with drug dependence in this study. A systematic review published in 2021 showed that substance use is a predictor for BD and (hypo)manic symptoms (Lalli et al., 2021). Specific data regarding cocaine use and BD has also been published. A prospective study investigated lifetime cocaine use as a potential predictor for conversion from major depressive disorder to bipolar disorder (de Azevedo Cardoso et al., 2020). The study analysis showed that the risk for conversion from major depressive disorder to BD was 3.41-fold higher in subjects who reported lifetime cocaine use at baseline. A systematic review also found a five-fold increased risk on the development of BD in individuals with lifetime cocaine use (Marangoni et al., 2016). Therefore, we consider this finding as part of the advancement of studies in the area, corroborating the information already established in the literature.

This study has some positive points to be noted. Initially, our sample is composed of young adults between 18 and 24 years old. According to the literature, BD symptoms usually appear before the age of 25, so the population used to build the model allows us to think about an early identification of the disease. This is possible because the population used to build the model was in a critical period of development for the onset of symptoms. Additionally, our team had an external psychiatrist to confirm the diagnosis whenever there were doubts through the standardized diagnostic interview (MINI), which sets a gold standard for characterization of the diagnosis. The average period between the initial interview and the follow-up was an average of five years, higher than in other studies in this field (Ribeiro et al., 2020).

The study has a large sample, collected through a probabilistic sample, obtained from the population of a city in southern Brazil with approximately 343,651 inhabitants. These factors bring robustness to our model. However, the outcome presented is difficult to predict due to: 1) the rarity of the outcome and 2) control participants may develop BD later. Nonetheless, this is a common challenge in studies in this area and we try to address these issues, whenever possible, statistically. Another point that must be taken into account when understanding the results presented here is the generalizability and applicability of the model. Studies in the area of precision psychiatry are on the rise. In this work, we aim and manage to present satisfactory results (test AUC 0.78), however, we understand that the data presented are primarily for scientific purposes and as a basis for future improvements. This study demonstrates that, in the near future, it will be possible to think of a calculator capable of being implemented in basic health systems. The information presented may be useful especially for patients who present characteristics seen here as of potential importance in the face of the diagnosis of BD: current depressive episode, depressive symptoms (mainly related to feelings of failure and sadness) and lifetime use of cocaine. Such a tool has the potential for robust screening, enabling symptomatic treatment, ensuring a better prognosis and preventing more severe clinical conditions.

In summary, we developed a binary model with a state-of-the-art algorithm capable of predicting the diagnosis of BD in approximately five years in a specific population of young adults, through clinical, socio-environmental, substance use, sex-related variables and demographic data, collected through a probabilistic sample. However, aiming for a better characterization of the BD diagnosis, future studies should focus on making systematic follow-ups that seek to follow these subjects during other developmental stages, as well as investing in studies that use specific risk populations, such as depressed patients or children of parents with BD. Furthermore, the inclusion of digital health data, biological and neuropsychological information and the use of neuroimaging can help in the rise of new models with greater applicability for the future.

## Data Availability

Research data are not shared due to privacy and anonymity reasons outlined by the authors of the original research project.

## Footnotes

1 Class decision boundary separates the data points into classes, where the algorithm switches from one class to another. In the present paper, a threshold of 0.5 was used. If a prediction had a probability *≥* 0.5, it was classified as a positive instance, otherwise, as a negative one.

